# Connectome dysfunction in patients at clinical high risk for psychosis and modulation by oxytocin

**DOI:** 10.1101/2023.03.17.23286528

**Authors:** Cathy Davies, Daniel Martins, Ottavia Dipasquale, Robert A. McCutcheon, Andrea De Micheli, Valentina Ramella-Cravaro, Umberto Provenzani, Grazia Rutigliano, Marco Cappucciati, Dominic Oliver, Steve Williams, Fernando Zelaya, Paul Allen, Silvia Murguia, David Taylor, Sukhi Shergill, Paul Morrison, Philip McGuire, Yannis Paloyelis, Paolo Fusar-Poli

**Author notes:** **Corresponding author:** Dr Cathy Davies, Department of Neuroimaging, Institute of Psychiatry, Psychology & Neuroscience, King’s College London, DeCrespigny Park, London, UK, SE5 8AF. These authors contributed equally to this work (Cathy Davies, Daniel Martins). These authors share senior authorship (Yannis Paloyelis, Paolo Fusar-Poli).

## Abstract

Abnormalities in functional brain networks (functional connectome) are increasingly implicated in people at Clinical High Risk for Psychosis (CHR-P). Intranasal oxytocin, a potential novel treatment for the CHR-P state, modulates network topology in healthy individuals. However, its connectomic effects in people at CHR-P remain unknown. Forty-seven men (30 CHR-P and 17 healthy controls) received acute challenges of both intranasal oxytocin 40 IU and placebo in two parallel randomised, double-blind, placebo-controlled cross-over studies. Multi-echo resting-state fMRI data was acquired at approximately 1h post-dosing. Using a graph theoretical approach, the effects of group (CHR-P vs healthy control), treatment (oxytocin vs placebo) and respective interactions were tested on graph metrics describing the topology of the functional connectome. Group effects were observed in 12 regions (all p_FDR_<.05) most localised to the frontoparietal network. Treatment effects were found in 7 regions (all p_FDR_<.05) predominantly within the ventral attention network. Our major finding was that many effects of oxytocin on network topology differ across CHR-P and healthy individuals, with significant interaction effects observed in numerous subcortical regions strongly implicated in psychosis onset, such as the thalamus, pallidum and nucleus accumbens, and cortical regions which localised primarily to the default mode network (12 regions, all p_FDR_<.05). Our findings provide new insights on aberrant functional brain network organisation associated with psychosis risk and demonstrate, for the first time, that oxytocin modulates network topology in brain regions implicated in the pathophysiology of psychosis in a clinical status (CHR-P vs healthy control) specific manner. Further profiling of the connectomic, clinical and cognitive effects of oxytocin in this population is warranted.

## INTRODUCTION

Abnormal functional connectivity is one of the strongest biological markers associated with psychosis (1–3). Alterations in the brain’s functional network organisation (functional connectome) are also observed prior to the onset of psychosis in people at Clinical High Risk (CHR-P) (4–6). These help-seeking individuals present with attenuated psychotic symptoms, emotional, cognitive and functional impairments and have a 20% two-year risk of transitioning to frank psychosis (7,8). There are currently no licensed pharmacological treatments for CHR-P patients (9,10), which represents a significant unmet clinical need. A deeper understanding of the connectomic abnormalities contributing to psychosis risk, and the potential ameliorative effects of experimental therapeutics, is urgently required.

Accumulating evidence from different imaging modalities has revealed brain-wide as well as regional dysfunction in people at CHR-P (11–13) and those with established psychosis (for reviews see (14,15)), particularly in the hippocampus, striatum, thalamus and frontal cortex— core components of the most influential circuit models of psychosis pathophysiology (16–18). Evidence of aberrant resting-state functional connectivity between brain regions (19–21) subsequently highlighted that psychosis-related dysfunction cannot entirely be described by spatially discrete differences in neural activation. Psychosis-related dysconnectivity can be identified both within and between large-scale networks, particularly involving frontoparietal, default mode and salience networks (22–24). This suggests that network-based approaches are needed to fully capture the complete spectrum of brain dysfunction associated with psychosis.

More recently, the application of graph theory (25) to neuroimaging data opened up new possibilities for exploring aberrant functional brain network organisation from micro-to-macro-scale levels of analysis (26). Using graph theory, brain regions are represented by nodes, the functional connections between nodes represented by edges, and modules defined as communities of highly intra-connected nodes that have fewer inter-module connections (27). Like any complex network, the topology (organisational properties) of the human brain shapes its capacity for information transfer, influencing higher-order functions such as cognition as well as its vulnerability to insult (26,28,29). A disruption to the finely-tuned balance of integration vs segregation may lead to loss of high-fidelity information transfer, cognitive dysfunction and the psychotic phenomenology characteristic of the CHR-P state and frank psychosis (1,27,30). Supporting this hypothesis, evidence suggests that people with established psychosis have abnormalities across global and local (node- or module-level) topological properties (27), including reduced small-worldness (31), clustering (32), hubness (33) and modularity (34), as well as changes in global and local efficiency (2). However, the network dysfunction that precedes psychosis onset in the CHR-P state is less well characterised.

The few studies conducted to date have shown that abnormal modular organisation in CHR-P individuals at baseline is associated with a three-fold transition rate to frank psychosis (4). Further work demonstrates that CHR-P individuals who go on to transition (vs controls and non-transitions) have altered topological centrality in frontal and anterior cingulate regions (5), reduced global efficiency and clustering (6) (although global differences are not always found in this population (5,35)), regional changes in nodal efficiency correlating with symptom severity (6), and extensive reorganisations of network community structure across most of the large-scale resting-state networks (6). Together, these data suggest that further mapping of CHR-associated functional connectomic alterations—and how they respond to experimental therapeutics—would not only enrich understanding of the neurobiological mechanisms driving psychosis onset but may also illuminate novel treatment targets.

One potential novel treatment is the neuropeptide oxytocin, which has neurobehavioural effects in multiple domains that could be beneficial for CHR-P individuals. These include anxiolytic effects (36), modulation of social and emotional cognition (37–40), hypothalamic-pituitary-adrenal axis regulation (36) and parasympathetic modulation of the heart rhythm (41). In our previous CHR-P work, we demonstrated that oxytocin modulates frontal activation during mentalising (42), anterior cingulate neurochemistry (43) and resting cerebral blood flow in the hippocampus among numerous other regions (44). Independent evidence further suggests that oxytocin modulates connectivity within resting-state networks in healthy volunteers (45,46) and ‘normalises’ aberrant connectivity in several clinical populations, including patients with social anxiety (47), post-traumatic stress disorder (48) and autism (49). Moreover, a recent study in healthy males demonstrated that a single dose of oxytocin was sufficient to modulate local functional network topology, including in regions and networks implicated in the pathophysiology of psychosis (50). This raises the possibility that oxytocin may ameliorate the connectomic dysfunction present in CHR-P individuals. It is unclear, however, whether oxytocin will have similar connectomic effects in CHR-P patients as in healthy controls. Supporting the hypothesis that a differential effect may exist is our recent finding that oxytocin increases cardio-parasympathetic activity in CHR-P but not in healthy males (41).

To address this gap, in this study we took a data-driven approach combining multi-echo resting-state blood-oxygen-level-dependent (BOLD) functional magnetic resonance imaging (fMRI) with graph-theory modelling. We investigated differences in global and regional topology of the functional connectome related to (a) CHR-P status (CHR-P patients vs controls; group effects), (b) the main effects of intranasal oxytocin vs placebo (treatment effects), and (c) group x treatment interactions to identify clinical status-specific effects of oxytocin.

## SUBJECTS & METHODS

### Participants

Thirty male, help-seeking CHR-P individuals aged 18-35 were recruited from a specialist early detection service in London, UK (51). A CHR-P status was determined using the Comprehensive Assessment of At-Risk Mental States (CAARMS) 12/2006 criteria (52). Seventeen healthy male controls, aged 19-34, were recruited as part of a related study (see below) (50). Full inclusion and exclusion criteria are detailed in the Supplementary Methods. In both studies, subjects were asked to abstain from using recreational drugs for at least 1 week and alcohol for at least 24 hours prior to each session. Urine screening was conducted before each session. The study received Research Ethics approval (London Bridge Committee: 14/LO/1692 and King’s College London Committee: PNM/13/14-163) and all subjects gave written informed consent.

### Design, Materials, Procedure

CHR-P and control data were combined from two related studies which were collaboratively designed so that their data could be analysed together. The CHR-P study (ISRCTN48799530) used a randomised, double-blind, single-dose challenge of intranasal oxytocin versus placebo in a crossover design (one-week wash-out). Participants self-administered 40 IU intranasal oxytocin or placebo using a standard nasal spray, as per our in-house protocol following recommended guidelines (Supplementary Materials) (53). Participants were randomly allocated to a treatment order (oxytocin/placebo or placebo/oxytocin). Following drug administration, participants underwent a battery of MRI sequences which started in the morning period to minimise potential effects of diurnal variation in oxytocin or vasopressin (54). Healthy control data came from a related randomised, double-blind, single-dose triple-dummy crossover study (50), where participants received oxytocin/placebo via three administration routes (nasal spray, nebulizer and intravenous infusion) in one of two fixed sequences: either nebulizer/intravenous/spray, or spray/intravenous/nebulizer. In three out of four sessions only one route of administration contained the active drug; in the fourth session, all routes delivered placebo or saline. The dose administered intranasally was 40 IU. For this study, we only used data from the placebo and nasal spray arms to maximise comparability to the protocol used in the CHR-P sample. Therefore, dose, device for drug administration, scanner and time post-dosing were matched across studies.

### MRI Acquisition, Preprocessing and Denoising

All scans were conducted on a General Electric Discovery MR750 3 Tesla system (General Electric, Chicago, USA) using a 32-channel head coil. The 8 min 10 s multi-echo resting-state fMRI scan was obtained starting at (M±SD) 62.6±3.1 min post-dosing in the CHR-P group and 57.01±3.38 min in healthy controls. Acquisition parameters, image preprocessing and denoising procedures (performed using the AFNI (55) tool meica.py (56,57)) are detailed in the Supplementary Methods.

### Functional Connectivity

Figure 1 provides an overview of the network analysis steps. To generate brain-wide functional connectivity matrices for each subject and treatment condition, we first extracted the BOLD time courses for each of the 66 cortical anatomical regions-of-interest (ROIs) included in the Desikan-Killiany atlas (58), with the addition of 9 bilateral subcortical regions (cerebellum, thalamus, caudate, putamen, pallidum, hippocampus, amygdala, accumbens, ventral diencephalon) and brainstem from the Harvard-Oxford Atlas, for each participant/session using *fslmeants* from the FMRIB Software Library (FSL). The bilateral frontal pole had poor coverage in our data and thus these 2 nodes were removed, leaving 83 nodes. We generated bivariate Pearson’s correlation matrices for the time courses of all possible pairs of ROIs (83×83) using Matlab/R2015b. Finally, we normalised our correlation measures to z-scores using the Fisher r-z transform. To ensure that any graph metric results were not driven by patient-control differences in overall connectivity strength (59), we examined mean functional connectivity, calculated as the average of the elements in the lower triangular matrix for each subject/condition.

**FIGURE 1.**
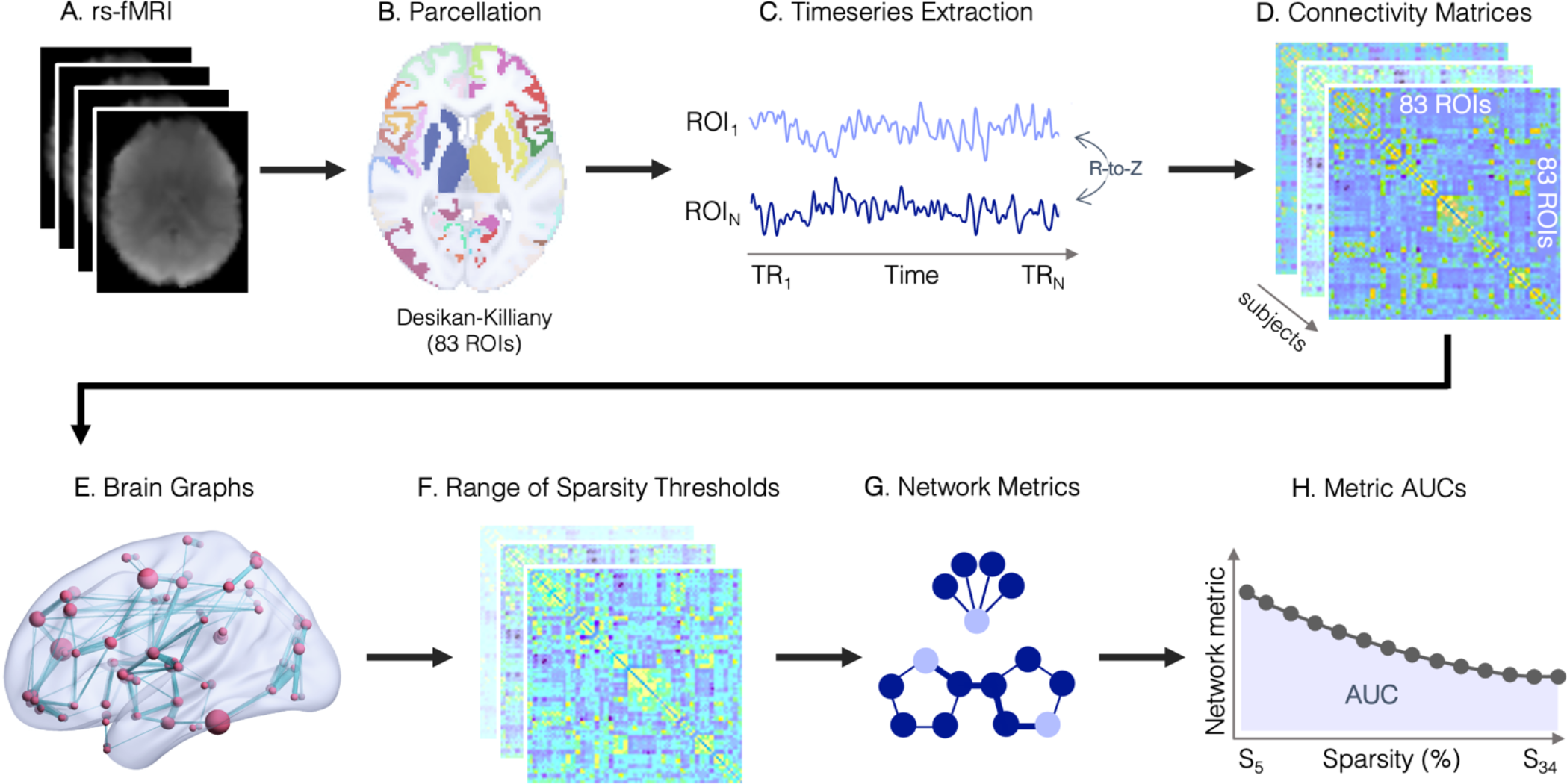
Overview of the network analysis steps.

### Graph Estimation and Network Characterisation

Our z-transformed connectivity matrices were then used to construct brain graphs representing the functional connectome for each subject/condition, using an undirected signed weighted approach (28). From the fully weighted networks we calculated equi-sparse networks by retaining a fixed percentage (K) of the total amount of possible edges. In order to determine systematic differences in the network’s topological organisation that are not dependent on the choice of an arbitrary threshold, we chose a range of sparsity thresholds from 5 to 34%, with steps of 1%, based on previous evidence showing the characteristic small-world behaviour of human brain networks is most consistently observed for this range (60). For each brain network and sparsity level, we used the Matlab functions provided with the Brain Connectivity Toolbox (28) to compute one global (global efficiency) and three nodal network metrics: betweenness-centrality, local efficiency and node degree. Details on the calculation of each of these metrics have been described elsewhere (28) but a brief overview is provided in Supplementary Figure S1. For each of the graph metrics analysed, we summarised the different values over the range of thresholds using the area under the curve (AUC), providing a summary estimator for each graph metric that is independent of single threshold selection and which is sensitive to topological differences in brain networks (61,62). All of our statistical analyses on graph metrics were therefore run on AUC parameters rather than the raw values.

### Statistical Analysis

We investigated group differences, treatment effects and (group x treatment) interactions across micro-to-macro-scale levels of functional connectome organisation. Specifically, we examined mean connectivity strength and global efficiency of the whole-brain network, the three nodal graph metrics, and individual connections between pairs of nodes. To evaluate the main effect of group (i.e. differences related to CHR-P status), treatment conditions were collapsed by calculating a mean average per subject and 2-sample t-tests conducted. For the main effect of treatment, oxytocin minus placebo (difference) values were calculated per subject and one-sample t-tests conducted. To examine interactions, the difference values were entered into two-sample (CHR-P vs control) t-tests. These analyses were conducted in Matlab and the directionality of significant interactions was determined by examining subject-level values.

#### Global Metrics

We calculated mean connectivity strength and global efficiency of the brain network for each participant and treatment condition and compared groups, treatments and interactions using the relevant t-tests, as above. Group differences in mean connectivity during the placebo condition (only) were also examined with a 2-sample t-test.

#### Node Level Metrics

We retrieved values for the betweenness-centrality, local efficiency and degree of each node of the connectome for each subject and treatment condition. Average and difference values were computed (as above) to examine the main effects of group, treatment and interaction effects using t-tests (as above), controlling false positives with FDR correction for the number of nodes examined.

#### Edge Level Metrics

To investigate differences in each individual edge of the connectivity matrices, we used the Network Based Statistics (NBS) method (63) implemented in the NBS toolbox v1.2 to control the family-wise error rate (in the weak sense) when mass-univariate testing is performed at every edge of the graph. While this approach does not allow for inferences on individual connections, it allows extraction of subnetworks or topological clusters of regions that are significantly differently connected between groups and conditions (63).

When compared to analyses at the individual edge level, the NBS method offers greater sensitivity while also controlling for false positives (63). In our NBS analysis, we tested for effects of group, treatment and interactions each using three arbitrary primary thresholds: 1.5, 3.1 and 4. We then used a two-tailed significance threshold of p<.05, with 5000 permutations.

For all analyses, statistical significance was set at p<.05 after correction for multiple testing, when applicable. Results were visualised using ENIGMA toolbox v2 (64), MRIcron and BrainNet Viewer. In the main text visualisation, all significant results were presented together on one brain template irrespective of the metric (betweenness-centrality, node degree and local efficiency), by assigning all significant t-statistics to their respective regions across metrics. Where there were effects in the same region in more than one metric, the larger t-value was shown. Further supplementary visualisations depicted results by individual metrics.

### Overlap with Large-Scale Resting-State Networks

To maximise the interpretability of our findings and facilitate comparisons with previous work focusing on large-scale resting-state networks (RSNs) (45,50), we quantified the percentage of overlap (Dice-kappa coefficient) between our group, treatment and interaction effect maps (separately) and each of the RSNs described in the atlas from Yeo et al (65), as detailed in the Supplementary Material and Fig 3. These values provide a qualitative contextualisation of our main findings which the reader can use for quick comparisons with previous literature.

**FIGURE 2.**
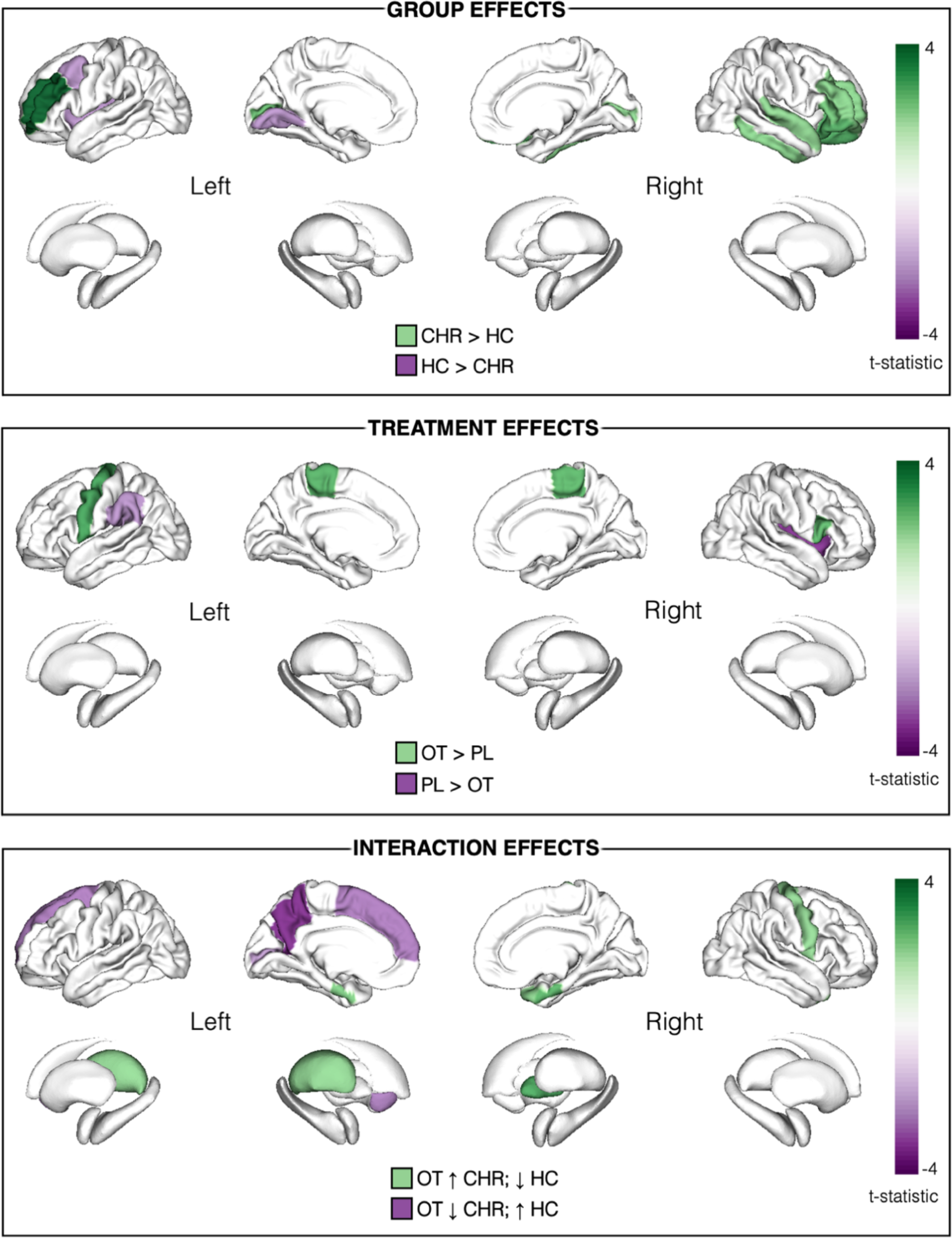
Overview of group, treatment and interaction effects across all nodal metrics (betweenness-centrality, node degree and local efficiency). The shade of colour represents the t-statistic for each region. Only regions surviving FDR-corrected significance threshold p<.05 shown. In the top panel depicting main effects of group, regions in purple and green depict lower and greater graph metrics (respectively) in CHR-P relative to healthy controls (HC). In the middle panel depicting treatment effects, regions in green and purple depict increases and decreases (respectively) in graph metrics under oxytocin (OT) relative to placebo (PL). Although significant, the brainstem (t= 2.34) is not depicted as it is not included in the ENIGMA visualisation template. In the lower panel depicting group x treatment interaction effects, regions in green depict where oxytocin increased (↑) graph metrics in the CHR-P group but decreased (↓) them in healthy controls; regions in purple depict where oxytocin decreased graph metrics in the CHR-P group but increased them in controls. All results are presented here irrespective of the metric; where there were effects in the same region in more than one nodal metric, the larger t-value is shown. All corresponding statistics are presented in Table 2 and individual figures for each metric are appended in Supplementary Figures 2-4.

**FIGURE 3.**
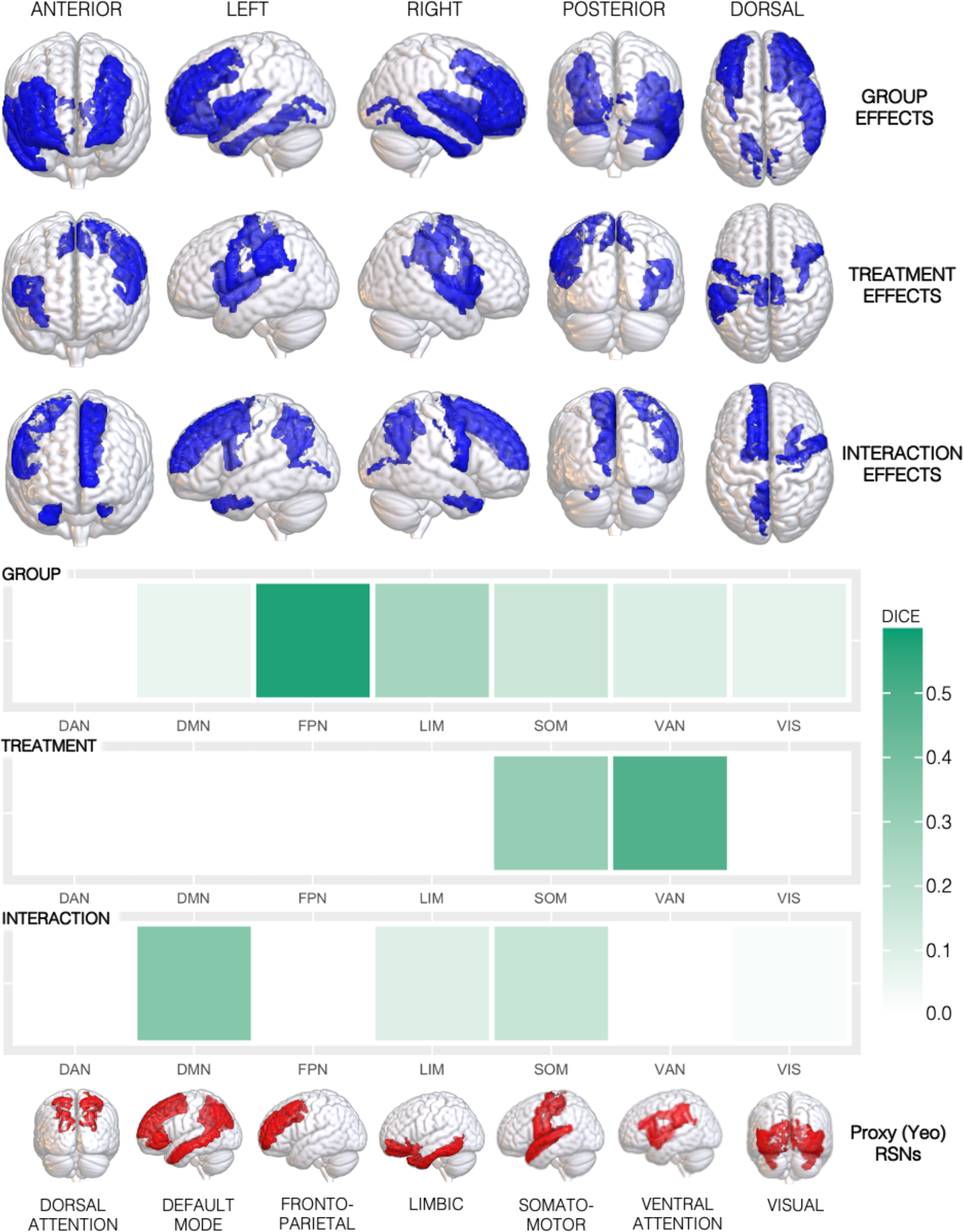
Overlap between the group, treatment and interaction findings and the large-scale canonical resting-state networks (RSNs). We calculated the percentage of overlap between our result maps—which included binary masks of all cortical regions showing differences in nodal metrics (for group, treatment and interaction effects, separately)—and the large-scale RSNs described in the atlas from Yeo et al. (65). As subcortical structures are not covered by the Yeo atlas, these were omitted from our result maps to prevent artificial reduction of the overlap estimate. The Yeo atlas includes a coarse parcellation of 7 canonical RSNs: the default-mode (DMN), dorsal attention (DAN), frontoparietal (FPN), limbic (LIM), somatomotor (SOM), visual (VIS) and ventral attention (VAN) networks. We created a proxy DKA>Yeo atlas for each of the 7 Yeo RSNs by combining individual DKA regions, allocating each to a single RSN based on the RSN for which each region had the highest number of overlapping vertices based on the confusion matrix from a previous study (100). Overlap was quantified using the Dice-kappa coefficient, which measures the percentage of voxels of each RSN overlapping with our group/treatment/interaction effect maps. In the upper section, we provide an overview of all regions where we found group, treatment or interaction effects, irrespective of the specific graph metric, rendered in a 3D glass brain (semi-transparent) surface model. In the lower section, we provide a heatmap summarising the percentage of overlap (Dice-kappa coefficient) between our results and each of the 7 networks, with each network rendered in a 3D glass brain (semi-transparent) surface model. Note that despite the visualisation, regions belonging to the different RSNs do not overlap, for example, the FPN contains the rostral and caudal middle frontal gyri, wheras the DMN contains the superior and inferior frontal gyri, which are difficult to differentiate in rendered models.

## RESULTS

### Sample Characteristics

Demographic and clinical characteristics of the sample are presented in Table 1. There were significantly more smokers in the CHR-P relative to the control group. One CHR-P subject was removed due to protocol violations and another omitted due to excessive head movement, leaving a sample of 28 CHR-P individuals and 17 controls.

**TABLE 1.** Participant Demographic and Clinical Characteristics.

| Variable | CHR-P (N=30) | Controls (N=17) | Statistics |
| --- | --- | --- | --- |
| Age, years; mean (SD) | 23.2 (4.7) | 24.2 (5.3) | $p = .51^c$ |
| Sex, N (%) male | 30 (100) | 17 (100) | N/A<sup>d</sup> |
| Ethnicity, N White/Black/Asian/Mixed | 16/6/4/4 | — | — |
| Handedness, N (%) right | 26 (87) | 17 (100) | $p = .15^e$ |
| Education, years; mean (SD) | 13.2 (1.9)<sup>g</sup> | —<sup>g</sup> | — |
| CHR-P Subtype<sup>a</sup>, N BLIPS/APS/GRD | 6/23/1 | N/A | N/A |
| CAARMS positive symptoms<sup>b</sup>; mean (SD) | 11.7 (3.3) | N/A | N/A |
| GF social score; mean (SD) | 6.8 (1.5) | N/A | N/A |
| GF role score; mean (SD) | 7.0 (1.7) | N/A | N/A |
| Current smoker, N (%) yes | 17 (57) | 4 (24) | $p = .028^f$ |
| Cigarettes/day; mean (SD) | 9.8 (6.0) | 4.5 (4.0) | $p = .12^c$ |
| Cannabis ever used, N (%) yes | 24 (80) | 10 (59) | $p = .18^e$ |
| Current alcohol use, N (%) yes | 26 (87) | 16 (94) | $p = .64^e$ |

<sup>a</sup> Comprehensive Assessment of At-Risk Mental States (CAARMS) subgroup, BLIPS Brief Limited Intermittent Psychotic Symptoms; APS Attenuated Psychotic Symptoms; GRD Genetic Risk and Deterioration. <sup>b</sup> Sum of the global (severity) ratings for positive subscale items (P1-P4) of the CAARMS. GF - Global Functioning (Role and Social) Scale. <sup>c</sup> independent t-test, <sup>d</sup> no statistical test necessary, <sup>e</sup> Fisher's Exact test, <sup>f</sup> chi-square test, <sup>g</sup> the majority of controls were university students (approximately between 13-16 years of education), whereas 43% of CHR-P were university students or had completed a degree - of the remaining CHR-P subjects, 27% were currently undertaking A-Levels/BTEC (11-13 years of education), 20% were in full-time or part-time work, and 10% were unemployed.

### Main Effect of Group (CHR-P vs Healthy Controls)

#### Global metrics

There was no main effect of group (t(43) = -0.86, p = .39), nor any difference between groups when comparing the placebo conditions alone (t(43) = -0.82, p = .42) on mean functional connectivity. There were no significant group effects (t(43) = -1.59, p = .12) on global efficiency.

#### Betweenness-centrality

Across conditions (oxytocin and placebo conditions combined), compared to healthy controls, CHR-P individuals had lower betweenness-centrality of the left caudal middle frontal gyrus and left insula, but greater betweenness-centrality of the right inferior and superior temporal gyri.

#### Nodal degree

CHR-P individuals had lower node degree of the left lingual gyrus compared to controls, but greater node degree of the right lateral orbitofrontal cortex and right pars triangularis.

#### Local efficiency

Compared to controls, CHR-P individuals had greater local efficiency of the bilateral pericalcarine cortex, bilateral rostral middle frontal gyrus and right pars orbitalis. There were no regions where local efficiency was significantly greater in controls vs the CHR-P group.

Details of significant nodal metric results are presented in Table 2 and Fig 2, with metric-level figures appended in Fig S2.

**TABLE 2.** Effects of group, treatment and interaction effects on nodal betweenness-centrality, degree and local efficiency.

| GROUP EFFECTS |  |  |  |
| --- | --- | --- | --- |
| Node | Direction | T statistic | P (FDR-corr) |
| **Betweenness-Centrality** |  |  |  |
| Caudal middle frontal gyrus, left | HC > CHR | -2.165 | 0.036 |
| Insula, left | HC > CHR | -2.236 | 0.031 |
| Inferior temporal gyrus, right | CHR > HC | 2.150 | 0.037 |
| Superior temporal gyrus, right | CHR > HC | 2.092 | 0.042 |
| **Node degree** |  |  |  |
| Lingual gyrus, left | HC > CHR | -2.217 | 0.032 |
| Lateral orbital frontal cortex, right | CHR > HC | 2.757 | 0.009 |
| Pars triangularis, right | CHR > HC | 2.353 | 0.023 |
| **Local Efficiency** |  |  |  |
| Pericalcarine cortex, left | CHR > HC | 2.451 | 0.018 |
| Rostral middle frontal gyrus, left | CHR > HC | 3.896 | <0.001 |
| Pars orbitalis, right | CHR > HC | 2.698 | 0.010 |
| Pericalcarine cortex, right | CHR > HC | 2.125 | 0.039 |
| Rostral middle frontal gyrus, right | CHR > HC | 2.142 | 0.038 |
| TREATMENT EFFECTS |  |  |  |
| Node | Direction | T statistic | P (FDR-corr) |
| **Betweenness-Centrality** |  |  |  |
| Brainstem | OT > PL | 2.337 | 0.024 |
| Supramarginal gyrus, left | PL > OT | -2.249 | 0.030 |
| Insula, right | PL > OT | -3.286 | 0.002 |
| **Node degree** |  |  |  |
| Precentral gyrus, left | OT > PL | 2.885 | 0.006 |
| **Local Efficiency** |  |  |  |
| Paracentral lobule, left | OT > PL | 2.699 | 0.010 |
| Paracentral lobule, right | OT > PL | 2.652 | 0.011 |
| Pars opercularis, right | OT > PL | 2.600 | 0.013 |
| INTERACTION EFFECTS |  |  |  |
| Node | Direction | T statistic* | P (FDR-corr) |
| **Betweenness-Centrality** |  |  |  |
| Precentral gyrus, right | OT > PL in CHR; OT < PL in HC | 2.062 | 0.045 |
| Thalamus, left | OT > PL in CHR; OT < PL in HC | 2.059 | 0.046 |
| Pallidum, right | OT > PL in CHR; OT < PL in HC | 2.785 | 0.008 |
| Precuneus cortex, left | OT < PL in CHR; OT > PL in HC | -3.387 | 0.002 |
| Superior frontal gyrus, left | OT < PL in CHR; OT > PL in HC | -2.561 | 0.014 |
| Accumbens, left | OT < PL in CHR; OT > PL in HC | -2.490 | 0.017 |
| **Node degree** |  |  |  |
| Precuneus cortex, left | OT < PL in CHR; OT > PL in HC | -2.437 | 0.019 |
| Superior frontal gyrus, left | OT < PL in CHR; OT > PL in HC | -2.139 | 0.038 |
| **Local Efficiency** |  |  |  |
| Entorhinal cortex, left | OT > PL in CHR; OT < PL in HC | 2.247 | 0.030 |
| Entorhinal cortex, right | OT > PL in CHR; OT < PL in HC | 2.633 | 0.012 |
| Temporal pole, right | OT > PL in CHR; OT < PL in HC | 2.102 | 0.041 |
| Pericalcarine cortex, left | OT < PL in CHR; OT > PL in HC | -2.061 | 0.045 |

**Footnotes:** FDR-corr indicates FDR-corrected P-values. \*Interaction t-test compared difference values (oxytocin minus placebo) between groups. OT, oxytocin; PL, placebo; CHR, Clinical High Risk for Psychosis; HC, healthy control.

#### Subnetworks

The NBS analyses did not identify any subnetworks where there were significant effects of group on connectivity after correcting for multiple testing, irrespective of the primary threshold used.

#### Overlap with RSNs

The pathways for which we identified significant nodal differences between CHR-P individuals and controls overlapped primarily with regions belonging to the frontoparietal network (Dice coefficient = 0.58) and to a lesser extent, the limbic network (Dice coefficient = 0.26). The overlap with other networks was numerically weaker. All overlap findings are displayed in Fig 3.

### Main Effect of Treatment (Oxytocin vs Placebo)

#### Global metrics

There were no significant treatment effects on mean functional connectivity (t(44) = 1.27, p = .21) nor global efficiency (t(44) = 0.44, p = .66).

#### Betweenness-centrality

Over all individuals (CHR-P and controls combined), compared to placebo, oxytocin increased the betweenness-centrality of the brainstem, while it decreased the betweenness-centrality of the left supramarginal gyrus and right insula.

#### Nodal degree

Compared to placebo, oxytocin increased node degree of the left precentral gyrus. There were no regions where oxytocin decreased node degree.

#### Local efficiency

Compared to placebo, oxytocin increased the local efficiency of the bilateral paracentral lobule and the right pars opercularis. There were no regions where oxytocin decreased local efficiency.

Details of significant nodal results are presented in Table 2, Fig 2 and Fig S3.

#### Subnetworks

The NBS analyses did not identify any subnetworks where there were significant treatment effects on connectivity after correcting for multiple testing, irrespective of the primary threshold used.

#### Overlap with RSNs

The pathways for which we identified significant nodal modulatory effects of oxytocin overlapped primarily with regions belonging to the ventral attention (Dice coefficient = 0.48) and somatomotor (Dice coefficient = 0.31) networks (Fig 3).

### Interaction Effects (Group x Treatment)

#### Global metrics

There were no significant interaction effects on mean functional connectivity (t(43) = -0.25, p = .80) nor global efficiency (t(43) = -0.85, p = .40).

#### Betweenness-centrality

Significant interaction effects were observed in a number of brain regions. In CHR-P individuals, compared to placebo, oxytocin increased the betweenness-centrality of the left thalamus, right pallidum and right precentral gyrus, whereas in controls, the betweenness-centrality in these regions was decreased after oxytocin. Conversely, in the left nucleus accumbens, left precuneus and left superior frontal gyrus, compared to placebo, oxytocin decreased betweenness-centrality in CHR-P individuals whereas it increased betweenness-centrality in controls.

#### Nodal degree

Two regions showed significant interaction effects. In CHR-P individuals, relative to placebo, oxytocin decreased node degree of the left precuneus cortex and left superior frontal gyrus, while node degree of these regions increased in controls after oxytocin vs placebo.

#### Local efficiency

Significant interaction effects were observed for four nodes. In CHR-P individuals, compared to placebo, oxytocin increased the local efficiency of the bilateral entorhinal cortex and right temporal pole. In each of these, oxytocin decreased the local efficiency in controls. The opposite pattern was observed in the left pericalcarine cortex, with oxytocin (vs placebo) decreasing the local efficiency in CHR-P individuals but increasing it in controls.

Details of significant nodal results are presented in Table 2, Fig 2 and Fig S4.

#### Subnetworks

The NBS analyses did not identify any subnetworks where there were significant interaction effects on connectivity after correcting for multiple testing, irrespective of the primary threshold used.

#### Overlap with RSNs

The pathways for which we identified significant nodal interaction effects overlapped primarily with regions belonging to the default-mode network (Dice coefficient = 0.35; Fig 3).

## DISCUSSION

Using a data-driven approach, we investigated differences in the topology of the functional connectome—at multiple levels of its hierarchical organisation—in people at CHR-P and examined whether intranasal oxytocin might attenuate some of these differences. We first demonstrated that (a) CHR-P individuals exhibit predominantly greater local graph metrics compared to healthy controls in regions most localised to the frontoparietal network, while normative global network efficiency is preserved, and (b) that oxytocin mainly produced increases in local graph metrics, predominantly in the ventral attention network. The major— and novel—finding of the current study was that many effects of oxytocin on network topology differ across CHR-P and healthy individuals, with significant interaction effects observed in numerous brain regions strongly implicated in psychosis onset, which localised primarily to the default mode network. Collectively, these findings provide new insights on alterations in functional network organisation associated with psychosis risk. Furthermore, they provide the first *in vivo* evidence that oxytocin modulates network topology in regions implicated in the pathophysiology of psychosis in a clinical status (CHR-P vs healthy control) specific manner, which strengthens the rationale for future studies investigating the therapeutic role of oxytocin in this clinical population.

### Differences in Network Topology Related to CHR-P Status

Our first finding was that most differences in local network topology related to CHR-P status (i.e. group effects) reflected greater nodal centrality in patients compared to controls. These effects mapped primarily to the frontoparietal resting state network, a major cognitive control system shown to be impaired in patients across the psychosis continuum (66,67) and transdiagnostic pathological conditions (68–70). Notably, we found greater local efficiency and nodal degree in CHR-P individuals across numerous frontal regions, indexing greater integration and importance of these nodes in the network, respectively (28). Despite the relatively few studies to have examined graph metrics in CHR-P cohorts to date, our results are supported by previous findings of increased toplogical centrality (5) and/or altered modular assignment (4,6) of these regions in CHR-P and established psychosis (33) samples.

Importantly, our group effects emerged against a backdrop of no differences in raw connectivity strength, suggesting that these nodal differences are instead signatures of a reorganisation of the brain’s functional network architecture (4,27). Whether these changes arise through proximal frontal dysfunction, or reflect compensatory changes in response to dysfunctional network dynamics occurring elsewhere (27), is unclear. Conceptually, however, such reorganisations of network architecture are thought to lead to inefficient information flow, aberrant input integration and manifestation of the psychotic and cognitive symptoms (1,27,30) characteristic of the CHR-P state, the latter of which may be particularly relevant given that half of our group differences were localised to the frontal cortex. Conversely, the lack of differences we observed in global efficiency suggests that *global* capacity for information transfer is preserved in our CHR-P sample. This is consistent with some (5) (but not all (6)) previous literature, including a large multi-cohort study using age-matched controls across the psychosis spectrum (35). Together, these findings lend support to the idea that the connectomic dysfunction preceding psychosis onset may be more subtle and characterised by changes in local functional network architecture (35).

### Effects of Oxytocin on Network Topology

We next showed that across all measures of local network topology, oxytocin predominantly increased graph metrics compared to placebo. Of particular interest was the finding that oxytocin increased betweenness-centrality in the brainstem. High betweenness-centrality reflects nodes that participate in a large number of shortest paths in the network, thereby acting as ‘bridging nodes’ that mediate information flow and network integration (28,71). Our findings accord with those in a previous overlapping study in healthy controls, where oxytocin increased the nodal degree—an index of the importance of a node, equal to its number of edges—also in the brainstem (50). The brainstem is considered a key autonomic hub in models of dopamino-oxytocinergic-mediated social and emotional cognition (72), is a purported substrate for salience detection (73) (through connections with the ventral tegmental area [VTA] and mesolimbic dopamine pathway (74)) and is one potential mediator of the tuning effects of oxytocin on these processes (73). Consistent with this, we found that oxytocin-induced changes in graph metrics mapped primarily onto the ventral attention network (65), an aggregate of the salience network (75) and the site of the strongest effects of oxytocin on timeseries connectivity in a previous report (46). Considering that our results were derived after collapsing across healthy controls and CHR-P individuals, effects of oxytocin within the ventral attention network, brainstem and other regions found here could be said to reflect common effects that operate irrespective of clinical status.

### Group x Treatment Interaction Effects

Our major—and to our knowledge, novel—finding was the presence of widespread interaction effects across all local graph metrics and across a multitude of brain regions, suggesting that oxytocin modulates network properties in a clinical status-specific manner. This is consistent with the differential effects of oxytocin on timeseries correlational connectivity reported in a variety of clinical populations vs healthy controls, including patients with social anxiety (47,76), post-traumatic stress disorder (48), and autism (49). Here we demonstrate, for the first time, that this is also the case in terms of effects on functional brain network topology in people at CHR-P.

Strikingly, oxytocin was found to have divergent effects on local topology in core regions implicated in the pathophysiology of psychosis, including betweenness-centrality of the thalamus and striatum (pallidum, nucleus accumbens [NAcc]), and local efficiency of the entorhinal cortex. In all of these regions except the NAcc, graph metrics were relatively lower in CHR-P individuals under placebo and were increased by oxytocin, with the reverse relationship seen in controls. The thalamus, hippocampus and striatum (including pallidum and NAcc) represent key components of the most influential circuit models of psychosis, which propose that hippocampal hyperactivation drives downstream striatal hyperdopaminergia via increased excitation of the NAcc, increased inhibition of ventral pallidum and thus reduced inhibition of midbrain/VTA dopamine neurons projecting to the NAcc and associative striatum (16–18,77). Given the well-established dysfunction within these regions in psychosis, one possibility is that oxytocin’s effects differ as a function of baseline network (dis)organisation. Nonpsychotic first-degree relatives have abnormal anatomic centrality in many of the regions reported here, including the pallidum, thalamus and hippocampus (78). Although we did not find significant effects in the hippocampus directly, the entorhinal cortex is part of the hippocampal formation and acts as a multilevel buffer, bi-directionally gating information flow between neocortex and hippocampus (79). Reduced nodal clustering has been observed in the hippocampus, thalamus and amygdala in CHR-P transitions vs non-transitions and controls (6), and adolescent-onset schizophrenia patients show decreased nodal efficiency and strength within highly integrative network hubs, most consistently in the hippocampal formation (2). Transition to psychosis is also associated with reduced thalamic efficiency (6) and thalamocortical dysconnectivity more broadly (21). If, as this evidence suggests, network topology is altered in CHR-P individuals, it is conceivable that the net effects of exogenously administered oxytocin on network organisation may well differ, even if its cognitive-behavioural effects are still geared towards optimised tuning of social salience/attentional orientation (80). Such interaction effects would echo those seen among the wider connectivity literature (81,82) where the greatest magnitude of the connectomic, cognitive and/or behavioural effects of oxytocin are often reported in those subjects with the greatest divergence (compared to controls) at baseline/under placebo (47,82).

Albeit in the context of interactions, in contrast to the other subcortical interaction effects we found relatively *greater* centrality of the NAcc in the CHR-P group under placebo, a region previously identified as a locus of CHR-associated structural rich-club dysfunction (83). The NAcc is also the terminus for dopamine neurons projecting from the midbrain/VTA (84) in the above-mentioned psychosis-linked circuit. Although speculative, our finding that in CHR-P patients, oxytocin decreased the centrality of the NAcc but increased centrality of the pallidum (which provides inhibition of the VTA) may suggest that oxytocin affects network properties— in mesolimbic dopamine pathway regions—in a psychosis risk-selective manner, and in a direction that ameliorates baseline dysfunction. Indeed, engagement of dopaminergic circuits seems to be a key locus for some of the effects of oxytocin (85) (for a review see (86)) and a wealth of evidence demonstrates interplay of the dopamine and oxytocin systems (87–92).

A further potential contributor to the differential effects of oxytocin is perturbations in the endogenous oxytocin system itself. Although direct and robust evidence for altered receptor number/distribution (and central/peripheral levels of oxytocin) is lacking, various stands of indirect evidence suggest that the oxytocin system is dysfunctional in psychosis (93) and is associated with symptom profiles (92), amygdala activation (94), susceptibility to psychosis, anhedonia-asociality and striatal-amygdala network connectivity (95). Overall, psychosis risk-related differences in network topology, dopamine function, the oxytocin system, as well as alterations in the set-points of other circuits are not mutually exclusive and may each contribute to the differential net effects of oxytocin on graph properties observed here.

### Limitations

A number of limitations warrant consideration. First, data from CHR-P and healthy control groups were from different studies that had slightly different experimental designs, with the controls participating in a triple-dummy study (using multiple administration methods on all occasions) and CHR-P subjects participating in a simple oxytocin vs placebo nasal spray challenge. While the two studies were conceived together specifically to ensure that their data could be combined for the present analyses, it is possible that residual effects from differing designs could underlie some of the group differences we observed. Second, a number of the clinical and demographic variables collected in the CHR-P group were not collected in controls. As such, we did not control for these potential confounders within the analyses. While there were no significant between-group differences for most key demographic factors, there were more smokers in the CHR-P relative to the control group. The within-subject element helped to mitigate the potential impact of this but we cannot rule out an effect on our between-group results. Our sample size was relatively large for pharmaco-MRI study including a CHR-P sample, but a recent paper on the reliability of graph analytic results suggested that sample sizes of more than 80 are needed (96). However, as well as collecting multi-echo data we used a rigorous denoising protocol with enhanced quality control procedures, a combination that has been shown to perform favourably in terms of signal-to-noise ratio compared to other denoising pipelines (57). We further sought to maximise reliability of our findings by using weighted graphs and metrics summarised (using AUC) across multiple cost functions. Finally, we excluded female subjects due to sexual dimorphism in oxytocinergic function (97,98) and studied only one (mid-to-high range) dose of oxytocin. In light of the now well-known dose-dependent neurophysiological properties of oxytocin (99), differential connectomic effects by dose and gender should be explored in future studies.

## Conclusions

Collectively, our findings provide new insights on aberrant functional brain network organisation associated with psychosis risk and demonstrate, for the first time, that oxytocin modulates network topology in brain regions implicated in the pathophysiology of psychosis in a clinical status-specific manner. Given the current lack of effective treatments for people at CHR-P, a deeper understanding of how functional connectomic alterations contribute to psychosis risk and onset, and the potential ameliorative effects of experimental therapeutics such as oxytocin, remain important avenues for future research.

## Data Availability

All data produced in the present study are not openly available.

## Acknowledgements

The authors wish to thank the study volunteers for their participation, members of the OASIS and THEDS clinical teams, and the radiographers at the Centre for Neuroimaging Sciences, King’s College London, who carried out the MRI scans.

## Author Contributions

Substantial contributions to conception and design (PFP, YP, PMc, SW, FZ, PA, SS), acquisition of data (CD, DM, AdM, VRC, UP, GR, MC, DO), analysis (CD, DM, OD) and interpretation of data (CD, DM, RAM), drafting of the article (CD, DM) or revising it critically for important intellectual content (all authors), study supervision (PFP, YP, PMc), acquisition of funding (PFP, YP, PMc, PMo, SS, DT, PA, FZ, SW), final approval of the version to be published (all authors).

## Supplementary Information

Supplementary Material is attached.

