## Supplementary Material for "Connectome dysfunction in patients at clinical high risk for psychosis and modulation by oxytocin"

**Supplementary Methods**

- Participants
- Design, Materials, Procedure
- Nasal Spray Administration
- MRI Acquisition
- Resting-State fMRI Preprocessing and Denoising
- Overlap with Large-Scale Resting-State Networks
- Definitions of Graph Metrics: Supplementary Figure S1

**Supplementary Results**

- Metric-Level Results: Supplementary Figures S2-S4

**References**

### SUPPLEMENTARY METHODS

#### Participants

Thirty male, help-seeking CHR-P individuals aged 18-35 were recruited from a specialist early detection service in London, UK (1). A CHR-P status was determined using the Comprehensive Assessment of At-Risk Mental States (CAARMS) 12/2006 criteria (2). Briefly, subjects met one or more of the following subgroup criteria: (a) attenuated psychotic symptoms, (b) brief limited intermittent psychotic symptoms (BLIPS, psychotic episode lasting <1 week, remitting without treatment), or (c) either schizotypal personality disorder or first-degree relative with psychosis (2), all coupled with functional decline. Individuals were excluded if there was a history of previous psychotic disorder (with the exception of BLIPS, some of whom may meet acute and transient psychotic disorder criteria (3)) or manic episode, exposure to antipsychotics, neurological disorder or current substance-use disorder, estimated IQ <70, acute intoxication on the day of scanning, and any contraindications to MRI or intranasal oxytocin or placebo. Seventeen healthy male controls, aged 19-34, were recruited as part of a related study (4). Controls were screened for psychiatric disorders using the Symptom Checklist-90-Revised (5) and Beck Depression Inventory-II (6) questionnaires, were medication free, had no history of drug abuse, tested negative on a urine screening test for recreational drugs, and consumed <28 units of alcohol per week and <5 cigarettes per day. In both studies, subjects were asked to abstain from using recreational drugs for at least 1 week and alcohol for at least 24 hours prior to each session. Urine screening was conducted before each session. The study received Research Ethics Service approval (14/LO/1692 and PNM/13/14-163) and all subjects gave written informed consent.

#### Design, Materials, Procedure

*CHR-P sample*

For descriptive purposes, we also collected information on medication history, use of alcohol, tobacco and cannabis, and functioning using the Global Functioning (GF) Role and Social scales (7).

*Healthy control sample*

The healthy control study employed a double-blind, placebo-controlled, triple-dummy, crossover design, where participants took part in 4 experimental sessions spaced 8.90 days apart on average (SD = 5.65, range: 5-28 days). In each session, participants received treatment via three different oxytocin/placebo administration routes, in one of two fixed sequences: either nebuliser/intravenous infusion/standard nasal spray, or standard nasal spray/intravenous infusion/nebuliser, according to the treatment administration scheme presented in Fig. 8 in (4). In 3 out of 4 sessions only one route of administration contained the active drug; in the fourth session, all routes delivered placebo or saline. Participants were randomly allocated to a treatment order (i.e. a specific plan regarding which route delivered the active drug in each experimental session) that was determined using a Latin square design. Unbeknown to the participants, the first treatment administration method in each session always contained placebo, while intranasal (spray or nebuliser) oxytocin was only delivered with the third treatment administration. This protocol maintained double-blinding while avoiding the potential washing-out of intranasally deposited oxytocin (as might be the case if oxytocin had been administered at the first treatment administration point and placebo at the third administration point). For the current analyses, only the placebo and spray datasets were included, to match the data collected in CHR-P participants.

**Nasal Spray Administration**

Participants followed our standard in-house protocol that we have used consistently across all of our studies and is consistent with the guidelines outlined by Guastella et al (8). Specifically, participants self-administered 10 puffs, each containing 0.1 ml Syntocinon (4 IU) or placebo, one puff every 30 s, alternating between nostrils (hence 40 IU oxytocin in total). Participants blocked the opposite nostril while administering each puff, and each puff was followed by an immediate, brief, sharp, snort.

**MRI Acquisition**

We aimed to collect resting-state fMRI data at approximately 60 mins post-dosing, in line with previous findings of the spatiotemporal profile of oxytocin-induced changes in cerebral blood flow, which demonstrated sustained effects over a ~20-73 minute period (post-intranasal administration) (9). The resting-state scan was obtained starting at (M±SD) 62.6 ± 3.1 min post-dosing in CHR-P and at 57.01 ± 3.38 min in controls. All scans were conducted on a General Electric Discovery MR750 3 Tesla system (General Electric, Chicago, USA) using a 32-channel head coil. During an 8 min 10 s scan, subjects were asked to lie with their eyes open looking at a centrally-placed fixation cross. Functional data was acquired using a single shot multi-echo echo planar imaging (EPI) sequence (TR = 2500 ms, TE's= 12, 28, 44 and 60 ms, FA= 80°, FOV= 240 mm, matrix= 64 x 64, slice thickness= 3 mm, 32 continuous descending axial slices, resolution = 3.75 × 3.75 × 3 mm, 192 volumes collected for each echo). A 3D high-spatial-resolution Inversion Recovery Spoiled Gradient Echo (IR-SPGR) T1-weighted scan was also acquired (TR/TE/TI = 7328/3024/400 ms; FA= 11°, FOV= 270 mm, slice thickness = 1.2 mm, slice gap = 1.2 mm, matrix = 256 × 256, resolution/voxel size = 1.1 × 1.1 × 1.2 mm).

**Resting-State fMRI Preprocessing and Denoising**

The multi-echo resting-state fMRI dataset was preprocessed using the AFNI (10) tool meica.py (11,12) and FMRIB Software Library (FSL). Preprocessing steps included volume re-alignment, skull stripping, time-series de-spiking and slice-time correction. Functional data were then optimally combined (OC) by taking a weighted summation of the first three echoes, using an exponential T2 weighting approach (13). While we acquired four echoes, we decided to combine only the first three echoes because unpublished work from our group has shown that the inclusion of the fourth echo can introduce artifacts due to its low signal-to-noise ratio. The OC data were then denoised with the Multi-Echo ICA approach implemented by the tool meica.py (11,12) to remove motion artifacts and other non-BOLD sources of noise. This denoising method has been shown to reduce non-BOLD sources of noise and increase the temporal signal-to-noise ratio more effectively than other standard regression approaches (11,12,14). White matter (WM) and cerebrospinal fluid (CSF) signals were first extracted from each participant's preprocessed datasets using standard eroded WM and CSF masks co-registered to each individual's space (15). Mean WM and CSF signals were then regressed out and a high-pass temporal filter with a cut-off frequency of 0.005 Hz applied.

We used the six rigid-body parameters extracted for each participant using AFNI to calculate the mean frame-wise displacement (FD). One CHR-P subject was removed from further analyses due to excessive head motion (FD>0.5), while all other participants' motion lied within an acceptable range (FD<0.5) for both sessions. A study-specific template representing the average T1-weighted anatomical image was built using the Advanced Normalization Tools (ANTs) toolbox (16). Finally, each participant's cleaned datasets were co-registered to its corresponding structural scan, then normalised to the study-specific template before warping to standard MNI152 space, with 2mm$^3$ resampling. The final normalised images were visually inspected to ensure the quality of the preprocessing and the absence of artifacts.

**Overlap with Large-Scale Resting-State Networks**

To maximise the interpretability of our findings and facilitate comparisons with previous work focusing on large-scale resting-state networks (RSNs) (4,17), we calculated the percentage of overlap between our result maps—which included binary masks of all cortical regions showing differences in nodal metrics (for group, treatment and interaction effects separately)—and the RSNs described in the atlas from Yeo et al (18). As subcortical structures are not covered by the Yeo atlas, these were omitted from our result maps to prevent artificial reduction of the overlap estimate. The Yeo atlas includes a coarse parcellation of seven canonical resting-state networks: the default-mode, dorsal attention, frontoparietal, limbic, somatomotor, visual and ventral attention networks. We created a proxy DKA>Yeo atlas for each of the 7 Yeo RSNs (18) by combining individual DKA regions, allocating each to a single RSN based on the RSN for which each region had the highest number of overlapping vertices based on the confusion matrix from a previous study (19). Overlap was quantified using the Dice-kappa coefficient, which estimates the percentage of voxels of each RSN that overlap with our group/treatment/interaction effect maps. These values provide a qualitative contextualisation of our main findings which the reader can use for quick comparisons with previous literature.

### Definitions of Graph Metrics

**Supplementary Figure S1.** Summary of the graph theory metrics used. Here we summarise the basic concepts (based on (4,20)) of the four graph-theory metrics used in the current study to investigate group (CHR-P vs control), treatment (oxytocin vs placebo) and interaction effects (group x treatment) on the functional connectome. Detailed information on metric calculations can be found in previous publications (20).

| Metric | Level | Concept | Schematic example |
|---|---|---|---|
| **Global efficiency** | Global | Average inverse shortest path length. Networks with low average shortest path lengths enable faster and more efficient information transfer. Therefore, those with greater *inverse* shortest path lengths can be thought of as better integrated | 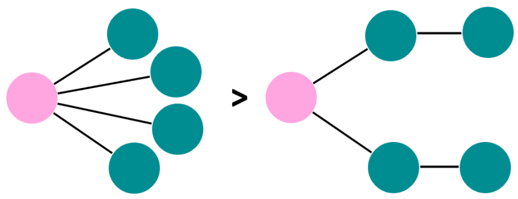 |
| **Local efficiency** | Local | Similar to global efficiency but the metric is computed on the neighbourhood of the given node. Nodes with high local efficiency can be thought of as having greater integration among its neighbours and better fault tolerance if the node is compromised | |
| **Betweenness-centrality** | Local | Fraction of all shortest paths in the network that pass through a given node. Nodes with high betweenness-centrality can be thought of as "bridging nodes" that connect different parts of the network | 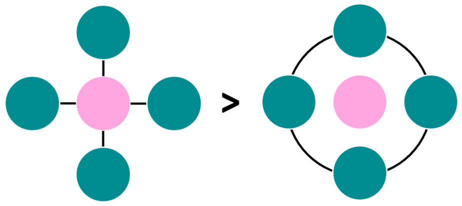 |
| **Node degree** | Local | Number of edges of a node. Nodes with high degree, i.e. with a large number of edges connecting it to other nodes, can be thought of as "hubs" | 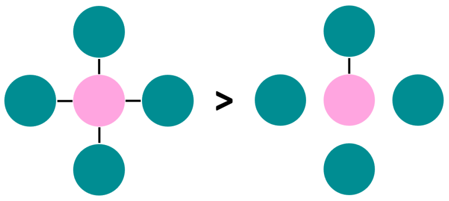 |

**Metric-level Results**

Results at the level of individual metrics (shown below) were visualised using BrainNet Viewer (21).

**Supplementary Figure S2.** Overview of group differences in betweenness-centrality, node degree and local efficiency. The pink and green node colours depict lower and greater graph metrics (respectively) in CHR-P relative to healthy controls (HC). The size of each nodal sphere is proportional to the T-statistic of each comparison (axial and sagittal sections scaled separately). ROI abbreviations: cMFG caudal middle frontal gyrus, STG superior temporal gyrus, ITG inferior temporal gyrus, Ling lingual gyrus, lOrbF lateral orbital frontal cortex, ParsT pars triangularis, rMFG rostral middle frontal gyrus, ParsOrb pars orbitalis, PCC pericalcarine cortex, r (suffix) right, l (suffix) left.

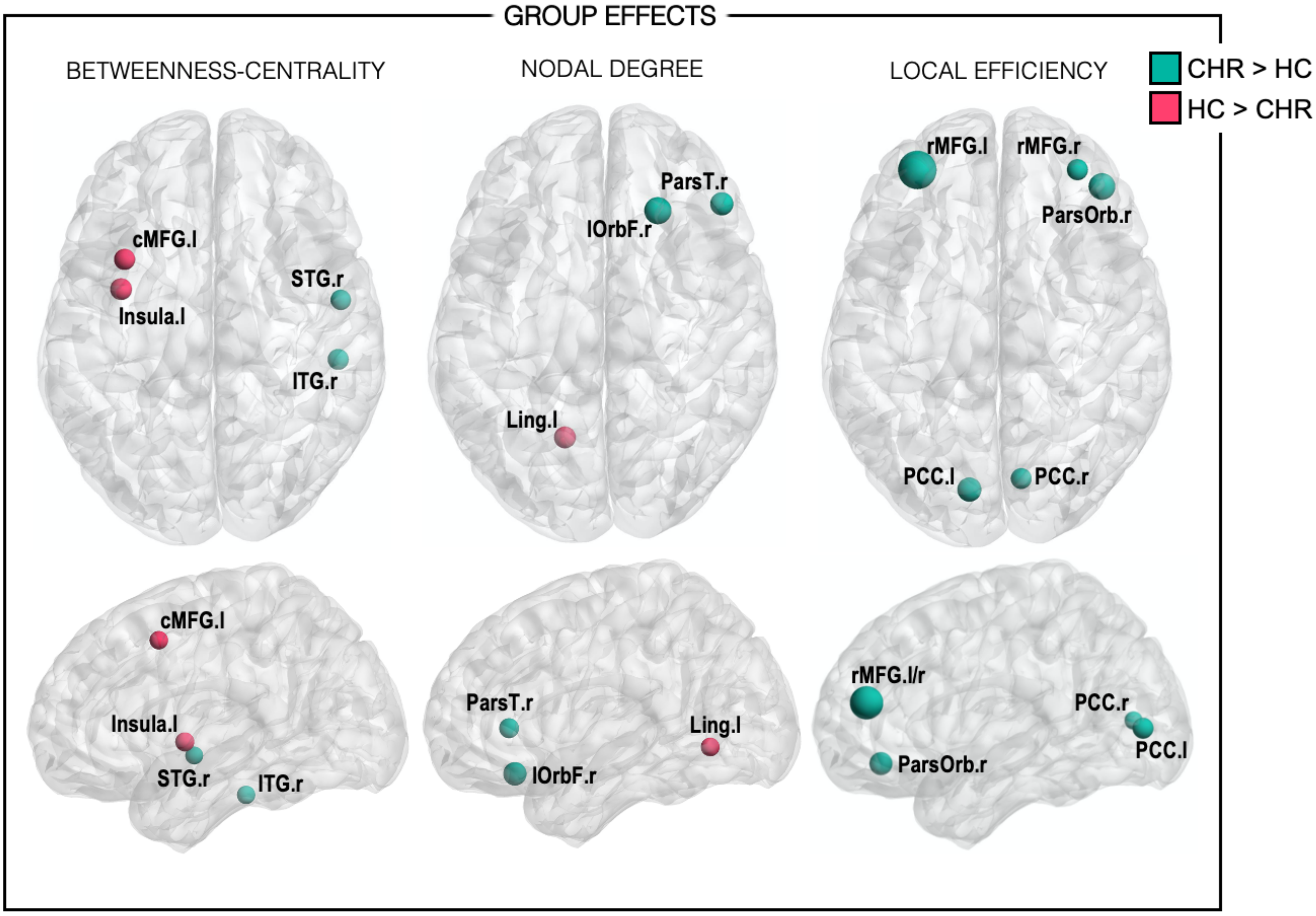

**Supplementary Figure S3.** Overview of oxytocin (treatment) effects on betweenness-centrality, node degree and local efficiency. The yellow and purple node colours depict increases and decreases (respectively) in graph metrics under oxytocin (OT) relative to placebo (PL). The size of each nodal sphere is proportional to the T-statistic of each comparison (axial and sagittal sections scaled separately). ROI abbreviations: SMG supramarginal gyrus, PrecG precentral gyrus, ParacL paracentral lobule, ParsOp pars opercularis, r (suffix) right, l (suffix) left.

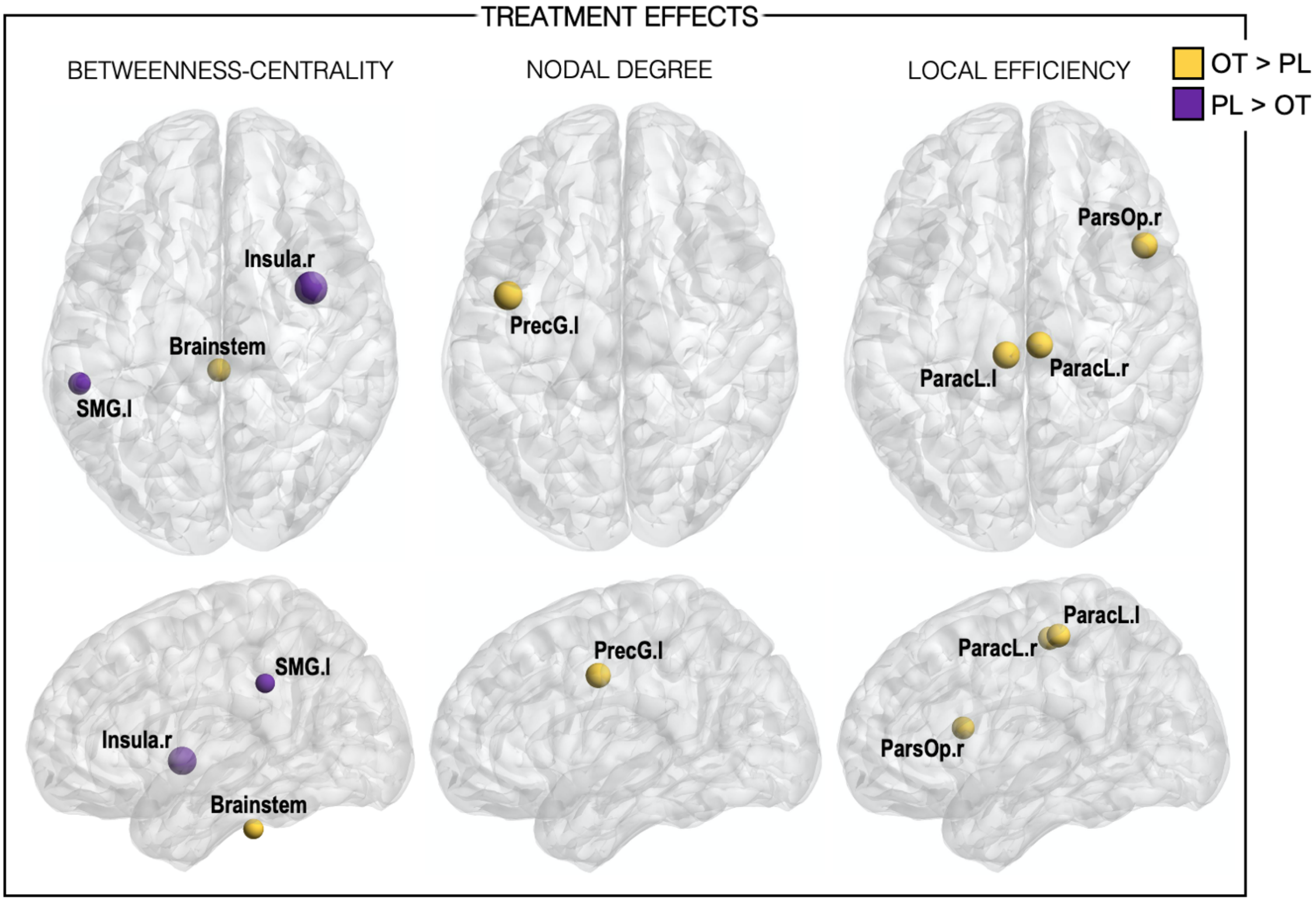

**Supplementary Figure S4.** Overview of group x treatment interaction effects on betweenness-centrality, node degree and local efficiency. The pink nodes depict where oxytocin (OT) increased (↑) graph metrics in the CHR-P group but decreased (↓) them in healthy controls (HC). The green nodes depict where oxytocin decreased graph metrics in the CHR-P group but increased them in controls. The size of each nodal sphere is proportional to the T-statistic of each comparison (axial and sagittal sections scaled separately). ROI abbreviations: SFG superior frontal gyrus, NAcc nucleus accumbens, Pall pallidum, PrecG precentral gyrus, Thal thalamus, Precun precuneus cortex, EHC entorhinal cortex, TP temporal pole, PCC pericalcarine cortex, r (suffix) right, l (suffix) left.

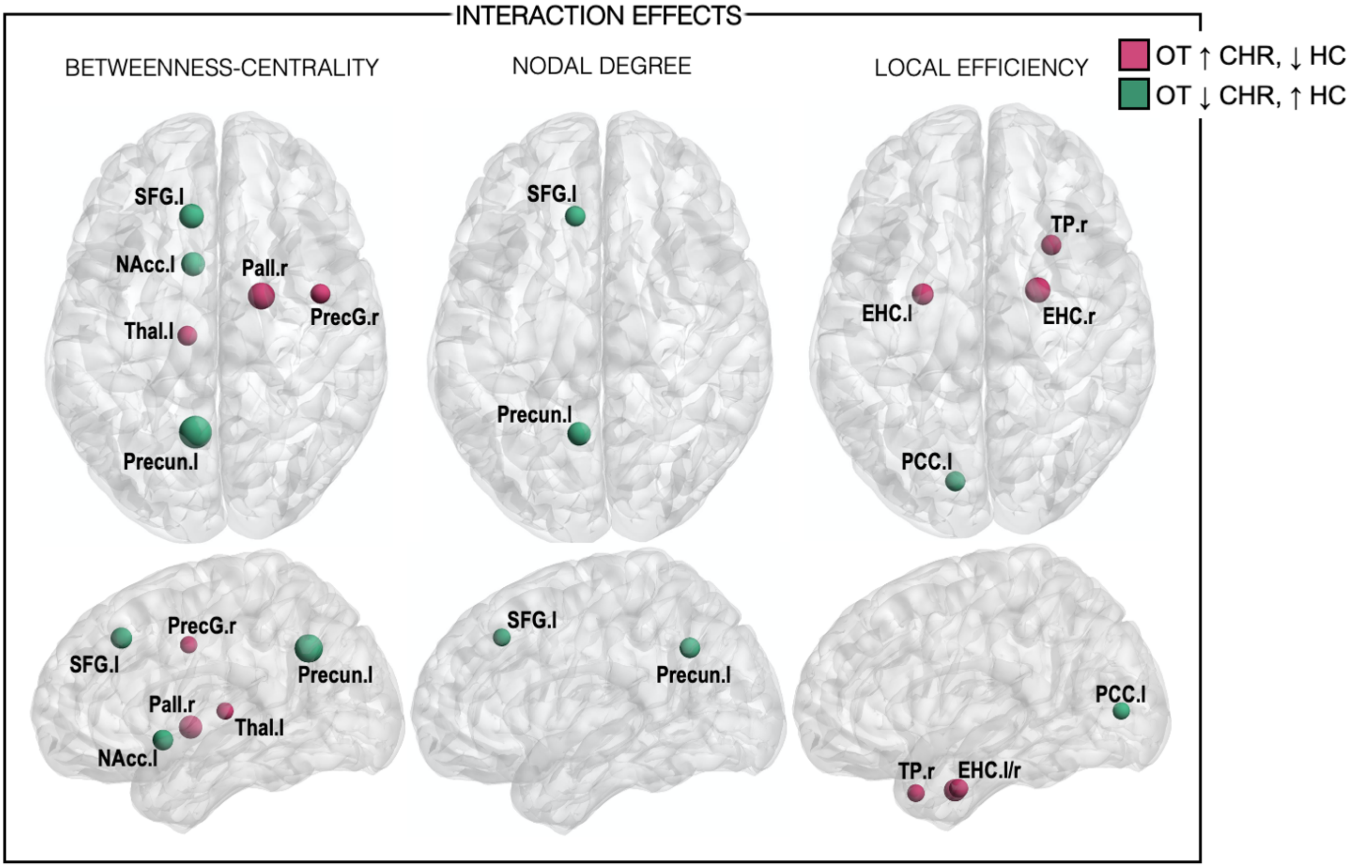
